# A Comparative Study of Predictive model (ECG Buddy) and ChatGPT-4o for Myocardial Infarction Diagnosis via ECG image Analysis: Performance, Accuracy, and Clinical Feasibility

**DOI:** 10.1101/2025.04.04.25325246

**Authors:** Haemin Lee, Sooyoung Yoo, Joonghee Kim, Youngjin Cho, Dongbum Suh, Keehyuck Lee

## Abstract

**Background:** Accurate and timely electrocardiogram (ECG) interpretation is critical for diagnosing myocardial infarction (MI) in emergency settings. Recent advances in multimodal Large Language Models (LLMs), such as Chat Generative Pre-trained Transformer (ChatGPT), have shown promise in clinical interpretation for medical imaging. However, whether these models analyze waveform patterns or simply rely on text cues remains unclear, underscoring the need for direct comparisons with dedicated ECG artificial intelligence (AI) tools.

**Methods:** This retrospective study evaluated and compared AI models for classifying MI using a publicly available 12-lead ECG dataset from Pakistan, categorizing cases into MI-positive (239 images) and MI-negative (689 images). ChatGPT (GPT-4o, version 2024-11-20) was queried with five MI confidence options, whereas ECG Buddy for Windows analyzed the images based on ST-elevation MI, acute coronary syndrome, and myocardial injury biomarkers.

**Results:** Among 928 ECG recordings (25.8% MI-positive), ChatGPT achieved an accuracy of 65.95% (95% confidence interval [CI]: 62.80–69.00), area under the curve (AUC) of 57.34% (95% CI: 53.44–61.24), sensitivity of 36.40% (95% CI: 30.30–42.85), and specificity of 76.20% (95% CI: 72.84–79.33). However, ECG Buddy reached an accuracy of 96.98% (95% CI: 95.67–97.99), AUC of 98.8% (95% CI: 98.3–99.43), sensitivity of 96.65% (95% CI: 93.51–98.54), and specificity of 97.10% (95% CI: 95.55–98.22). DeLong’s test confirmed that ECG Buddy significantly outperformed ChatGPT (all P < .001). In an error analysis of 40 cases, ChatGPT provided clinically plausible explanations in only 7.5% of cases, whereas 35% were partially correct, 40% were completely incorrect, and 17.5% received no meaningful explanation.

**Conclusion:** LLMs such as ChatGPT underperform relative to specialized tools such as ECG Buddy in ECG image-based MI diagnosis. Further training may improve ChatGPT; however, domain-specific AI remains essential for clinical accuracy. The high performance of ECG Buddy underscores the importance of specialized models for achieving reliable and robust diagnostic outcomes.

## Introduction

Electrocardiogram (ECG) interpretation is a fundamental skill in cardiovascular medicine, playing a crucial role in diagnosing conditions such as ST-elevation myocardial infarction (STEMI), arrhythmias, and electrolyte imbalances [1]. Accurate and timely ECG analysis is critical in clinical decision-making, particularly in emergency settings where rapid intervention can substantially impact patient outcomes. With advancements in artificial intelligence (AI), researchers have explored using various deep learning techniques—including convolutional neural networks and transformer-based models—to automate ECG interpretation by extracting clinically relevant features from ECG signal or image data [2–5].

Recently, multimodal Large Language Models (LLMs) trained on textual and imaging data have gained attention in the medical field [6]. These models have demonstrated the ability to generate diagnostic reports from chest radiographs, highlighting their potential for medical image interpretation [7]. As LLMs have become increasingly sophisticated, the interest in applying similar multimodal architectures to ECG interpretation has also grown.

General-purpose LLMs, such as OpenAI’s Generative Pre-trained Transformer (ChatGPT), have recently demonstrated promising capabilities in assisting with image interpretation and text-based medical assessments [8,9]. Unlike traditional AI models specifically trained for ECG signal processing, these models leverage extensive general knowledge and are now being considered for processing visual medical data, including ECG images. For example, Zaboli et al. [10] investigated the ECG interpretation ability and outcome prediction of ChatGPT in the emergency department and found moderate agreement with cardiologists, but with notable discrepancies in major adverse cardiac event (MACE) risk assessment. Zhu et al. [11] reported that GPT-4 achieved approximately 83% accuracy in multiple-choice ECG diagnostic questions, Günay et al. [12] ompared GPT-4, GPT-4o, and Gemini Advanced against cardiologists and emergency medicine specialists using routine and challenging ECG cases. Although all LLMs underperformed compared to cardiologists, GPT-4o showed relatively better accuracy and moderate agreement, suggesting potential as a supportive tool in clinical settings. Similarly, Avidan et al. [13] examined GPT-4o’s ability to detect atrial fibrillation in ECGs with confounding factors. Their findings indicated that while GPT-4o’s overall accuracy was comparable to that of internists and primary care physicians, it fell significantly short of cardiologists’ performance, particularly in challenging scenarios. In contrast, Günay et al. [14] reported that GPT-4 outperformed emergency medicine specialists in interpreting everyday ECG cases and performed on par with cardiologists when facing more complex ECG challenges.

Collectively, these studies highlight that although LLM-based approaches in ECG interpretation hold promise, their reliability in complex cases remains limited. Moreover, it remains unclear whether these models truly analyze waveform patterns or simply rely on text-based cues, such as machine-readable annotations. This raises concerns about the reproducibility of the models’ interpretations when presented with raw ECG images alone.

To date, no study has systematically compared the performance of LLMs against specialized ECG diagnostic AI tools. This comparison is becoming increasingly relevant, as general-purpose LLMs are not specifically designed for cardiovascular medicine. However, speculation about their potential applications in ECG interpretation is already widespread. Thus, a comparative evaluation with dedicated ECG AI software is necessary to determine the feasibility of LLM-based ECG interpretation in clinical practice.

Recent evaluations have demonstrated that ChatGPT-4o consistently outperforms other LLMs in ECG interpretation tasks, which makes it promising for comparative analyses [12]. It has been validated in multiple studies, demonstrating superior diagnostic accuracy to clinical experts in detecting conditions such as myocardial infarction (MI), hyperkalemia, and right ventricular (RV) dysfunction [15–18].

Therefore, in this study, we directly compared the performance of ChatGPT-4o with that of a commercially available dedicated AI-driven ECG analysis tool (ECG BuddyTM) in analyzing ECG images for MI detection. MI interpretation is one of the most essential aspects of ECG analysis.

Through this comparative study, we aimed to determine whether ChatGPT is currently capable of being effectively utilized for ECG interpretation in clinical practice.

## Materials and methods

### Study Design

In this retrospective study, we evaluated the performance of ChatGPT and ECG Buddy in classifying MI from ECG images. A publicly available 12-lead ECG image dataset compiled by the Ch. Pervaiz Elahi Institute of Cardiology in Multan, Pakistan, was used [19]. The dataset includes ECG images categorized into the following four groups: patients with MI (239 images), patients with abnormal heartbeats (233 images), patients with a history of MI (172 images), and healthy controls (284 images). The dataset does not provide additional patient information beyond these labels.

This study was designed and reported in accordance with the TRIPOD-LLM guidelines—a comprehensive reporting framework for studies involving LLMs in healthcare—to ensure that every step, from data processing and image-to-text conversion to AI querying and performance evaluation, was transparently and reproducibly documented [20]. To ensure consistency in data processing, extraneous areas of the images—including any supplementary text not related to patient information or diagnosis—were cropped, retaining only the waveform regions. For classification, only the images labeled as “myocardial infarction patients” were designated as MI-positive, representing active MI cases. The remaining 689 images—comprising abnormal heartbeats, history of MI, and healthy cases—were classified as MI-negative. The study design was approved by the Institutional Review Board of Seoul National University Bundang Hospital (IRBX-2504-966-902). Given the public availability of the dataset, the Institutional Review Board of Seoul National University Bundang Hospital granted a waiver for the requirement of informed consent

### AI Query and Output–ChatGPT

For the ChatGPT analysis, we used GPT-4o (version 2024-11-20) to process the identical set of ECG images. To assess the ability of ChatGPT to classify MI from ECG images, we designed a structured prompt aimed at systematically capturing the model’s diagnostic rationale and confidence in detecting MI. Before querying, ECG images were converted into base64 format, ensuring seamless integration with the ChatGPT application programming interface (API). ChatGPT was then provided with these base64-encoded images and prompted explicitly to analyze them, determine the likelihood of MI, and select from five predefined response categories—unknown, unlikely, possible, probable, and definite—representing increasing diagnostic certainty (Textbox 1). To define a positive MI diagnosis based on this likelihood measure, the Youden index was applied to determine an optimal cutoff. Specifically, ChatGPT was instructed to identify and specify which ECG leads exhibited abnormalities, describe the changes observed (such as ST-segment elevation, T-wave inversion, or pathological Q waves), and provide detailed reasoning supporting its diagnostic conclusion. In cases where no abnormalities were noted or the ECG findings were ambiguous, the model was prompted to discuss potential alternative diagnoses or clearly explain why the ECG appeared normal. All queries were conducted using GPT-4o via the ChatGPT API. An example of a typical ChatGPT response is illustrated in Figure 1.

**Fig 1.**
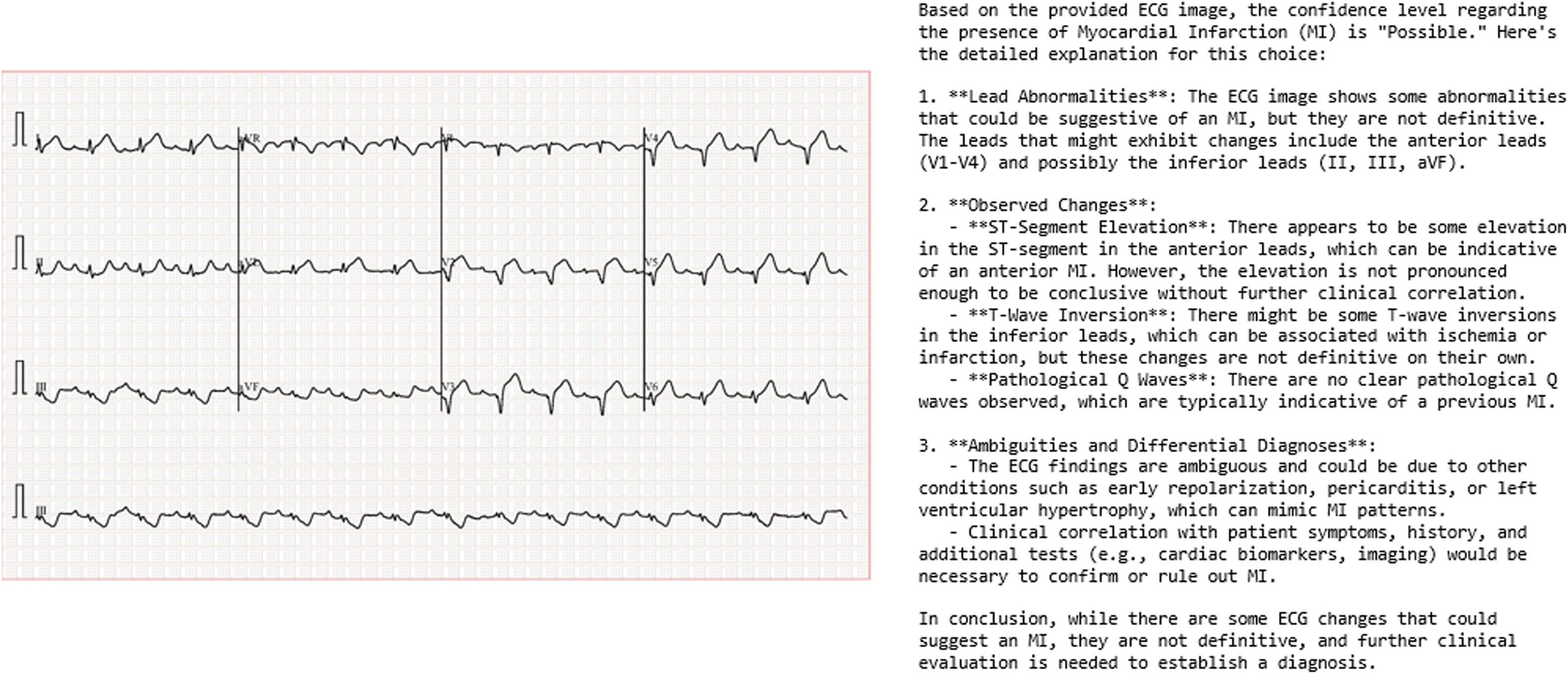
ECG Dataset and GPT output response.

### Qualitative Assessment of Diagnostic Explanations–ChatGPT

Additionally, a qualitative assessment of the diagnostic explanations of ChatGPT was performed to further evaluate the ability of ChatGPT to accurately interpret ECG images. A subset of 40 cases was randomly selected for detailed qualitative analysis. This subset comprised 10 true-positive, 10 true-negative, 10 false-positive, and 10 false-negative examples. A board-certified emergency physician with 14 years of experience then reviewed the diagnostic explanations of ChatGPT to assess whether the model provided clinically appropriate rationales that supported its classification decisions.

### AI-Powered Image Analysis–ECG Buddy

ECG Buddy is a deep learning-based ECG analysis platform designed for 12-lead ECG image interpretation. The software is available for both smartphones and Windows-based desktop personal computers. In this study, ECG Buddy for Windows was used to perform bulk analysis of ECG data (Figure 2). It is approved by the Korean Ministry of Food and Drug Safety (MFDS) and freely available for download in Korean appstores, can analyze 12-lead ECGs by taking pictures of ECG images to produce 10 digital biomarkers, The software automatically detects the ECG image displayed on the desktop and provides the analysis results within 10–15 s. Figure 2A shows the operating screen of ECG Buddy for Windows, while Figure 2B shows the ECG image analysis output. ECG Buddy generates 10 digital biomarkers that assess a range of cardiac conditions, including STEMI, acute coronary syndrome (ACS), myocardial injury (MyoInj), critical condition, pulmonary edema, pericardial effusion, left ventricular dysfunction, RV dysfunction, pulmonary hypertension, and severe hyperkalemia. This study analyzed only the STEMI, ACS, and MyoInj biomarkers owing to their direct relevance to MI classification.

**Fig 2.**
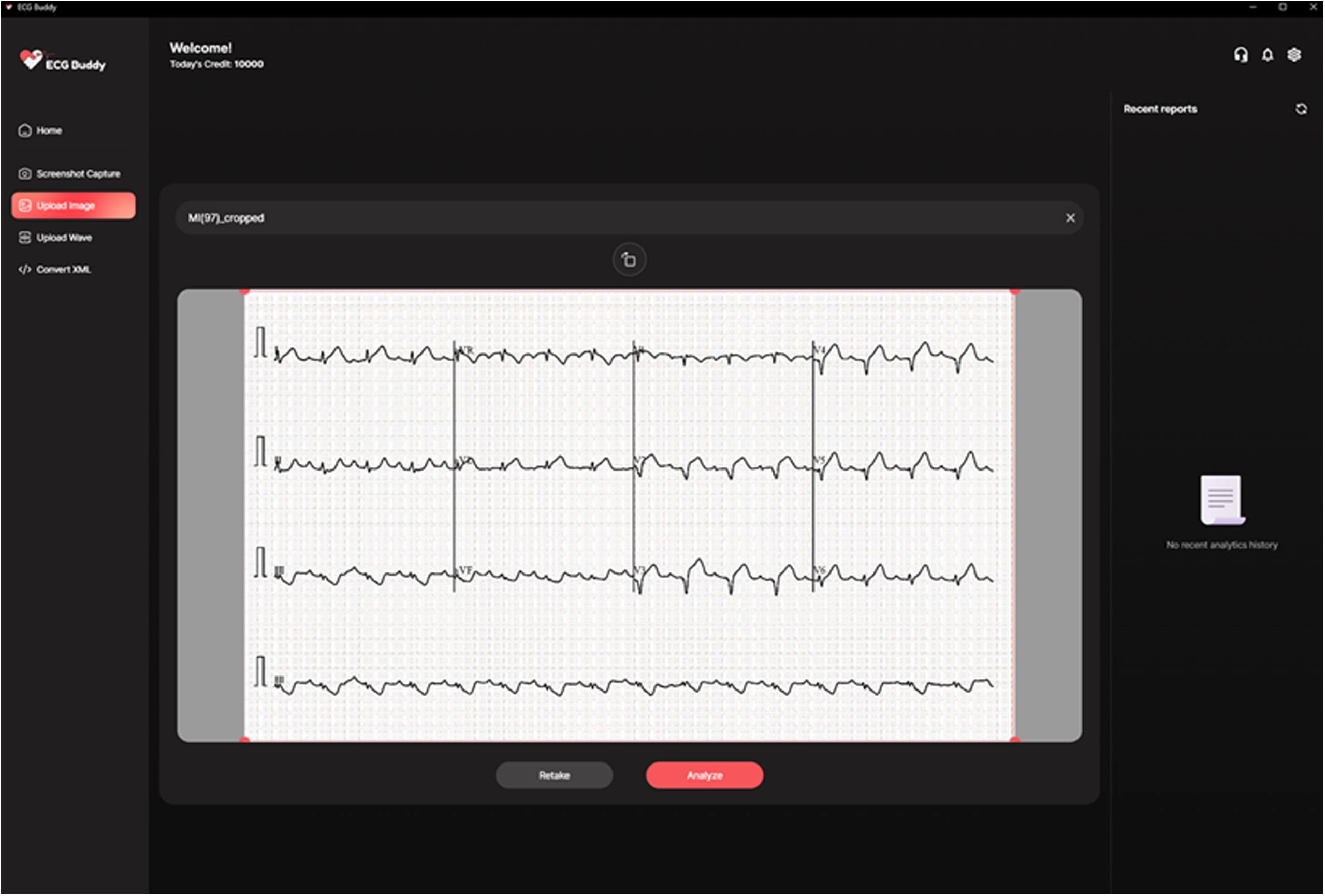
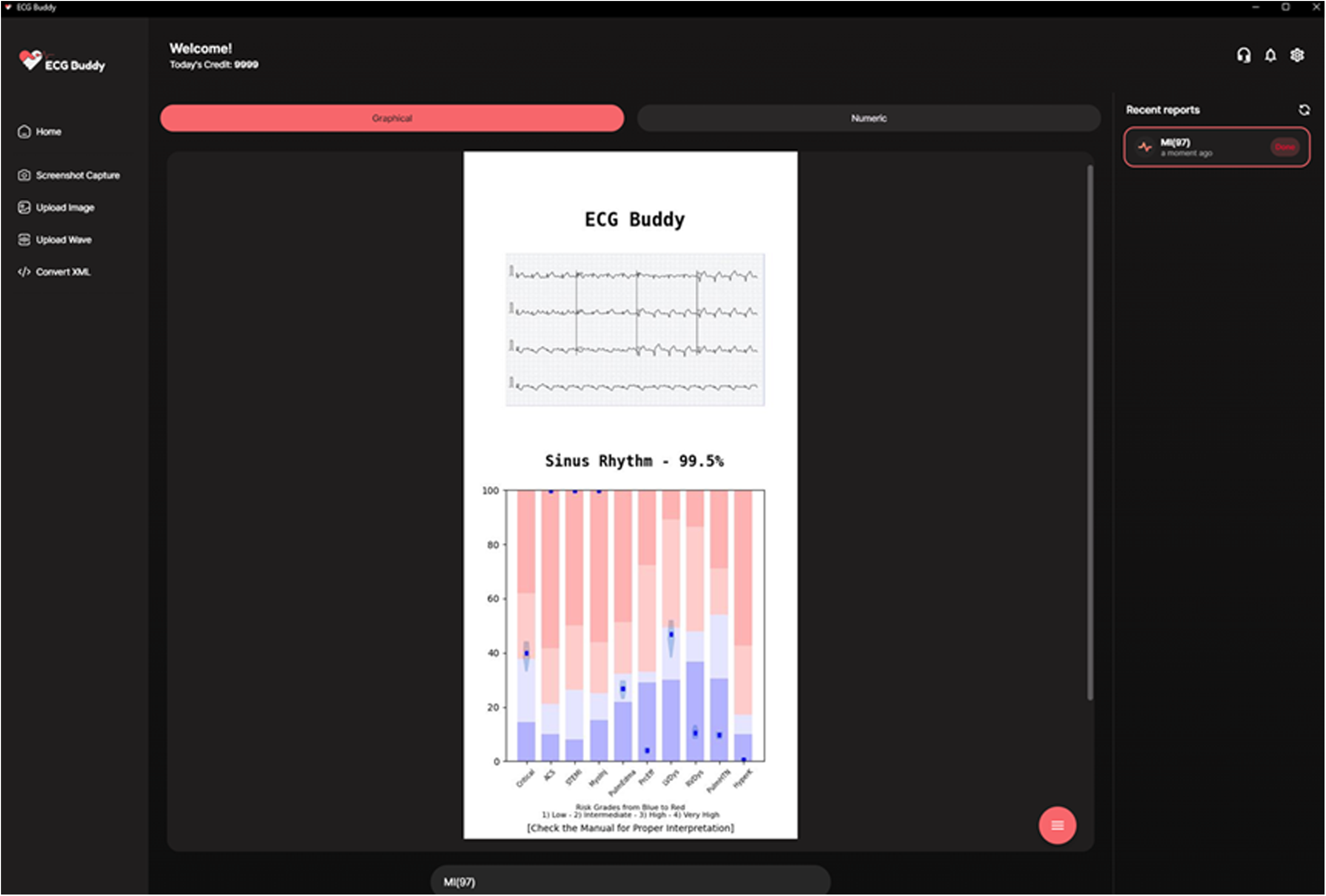
The operating screen of ECG Buddy for Windows. A: ECG input image, B: ECG image analysis result

### Statistical Analysis

Model performance was evaluated using accuracy, sensitivity, specificity, positive predictive value (PPV), and negative predictive value (NPV). The Youden index was used to determine optimal classification thresholds. Additionally, model performance was evaluated using the area under the receiver operating characteristic curve (AUC), and the AUC values were compared using DeLong’s method, with statistical significance set at P < .05. All analyses were conducted using R software version 4.1.0 [21], with ChatGPT API responses obtained using Python.

## Results

### Performance of ChatGPT and ECG Buddy

In total, 928 ECG recordings (25.8% MI-positive cases) were analyzed, and all were successfully processed by both AI models. ChatGPT demonstrated limited discriminative ability in MI detection, achieving an AUC of 57.34% (95% confidence interval [CI]: 53.44–61.24). Using the Youden index, the optimal cutoff was determined as the category “definite.” At this cutoff, the model’s sensitivity, specificity, PPV, and NPV were 36.40% (95% CI: 30.30–42.85), 76.20% (95% CI: 72.84–79.33), 34.66% (95% CI: 28.79–40.90), and 77.55% (95% CI: 74.21–80.64), respectively (Figure 3A and Table 1).

**Fig 3.**
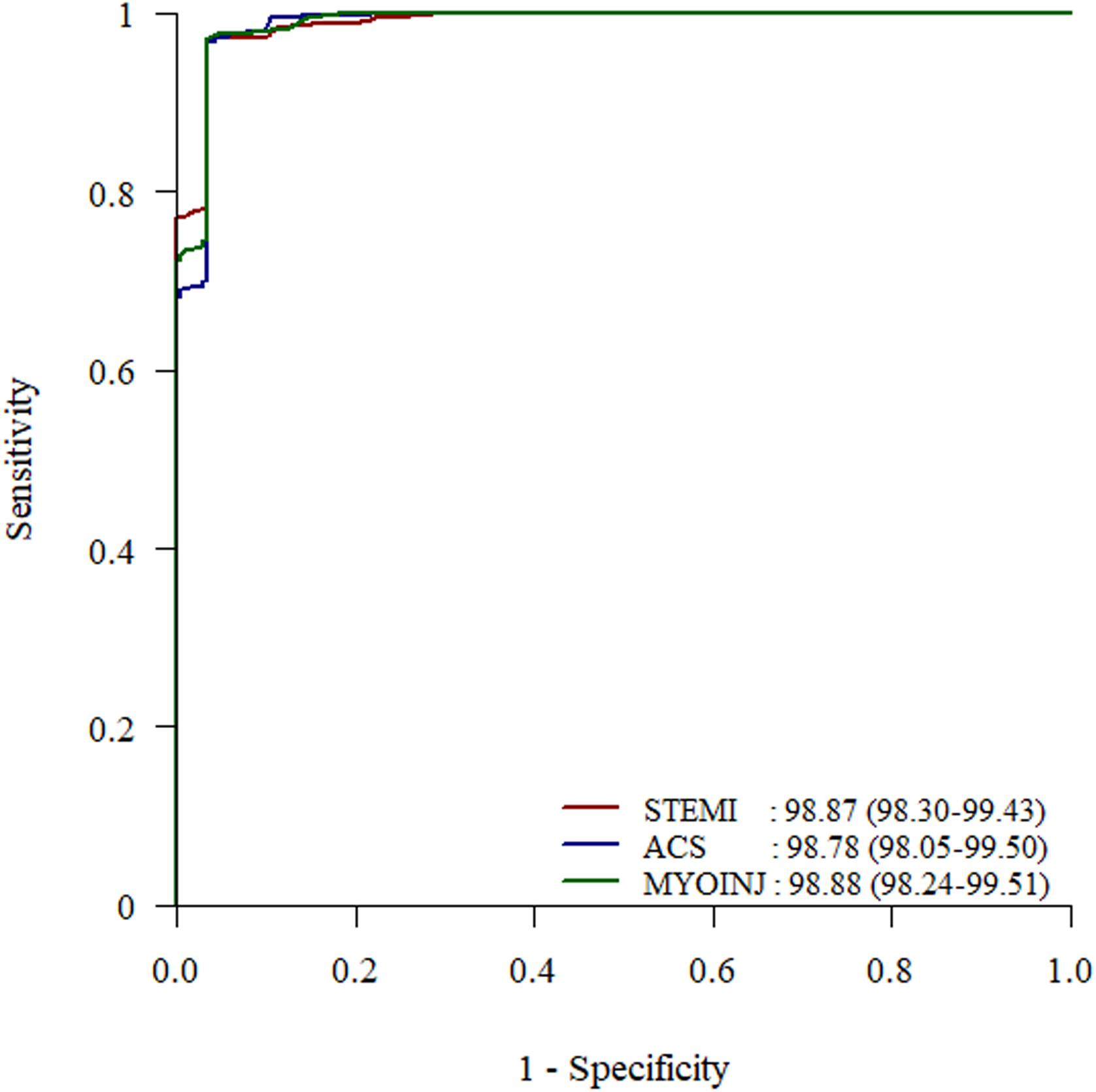
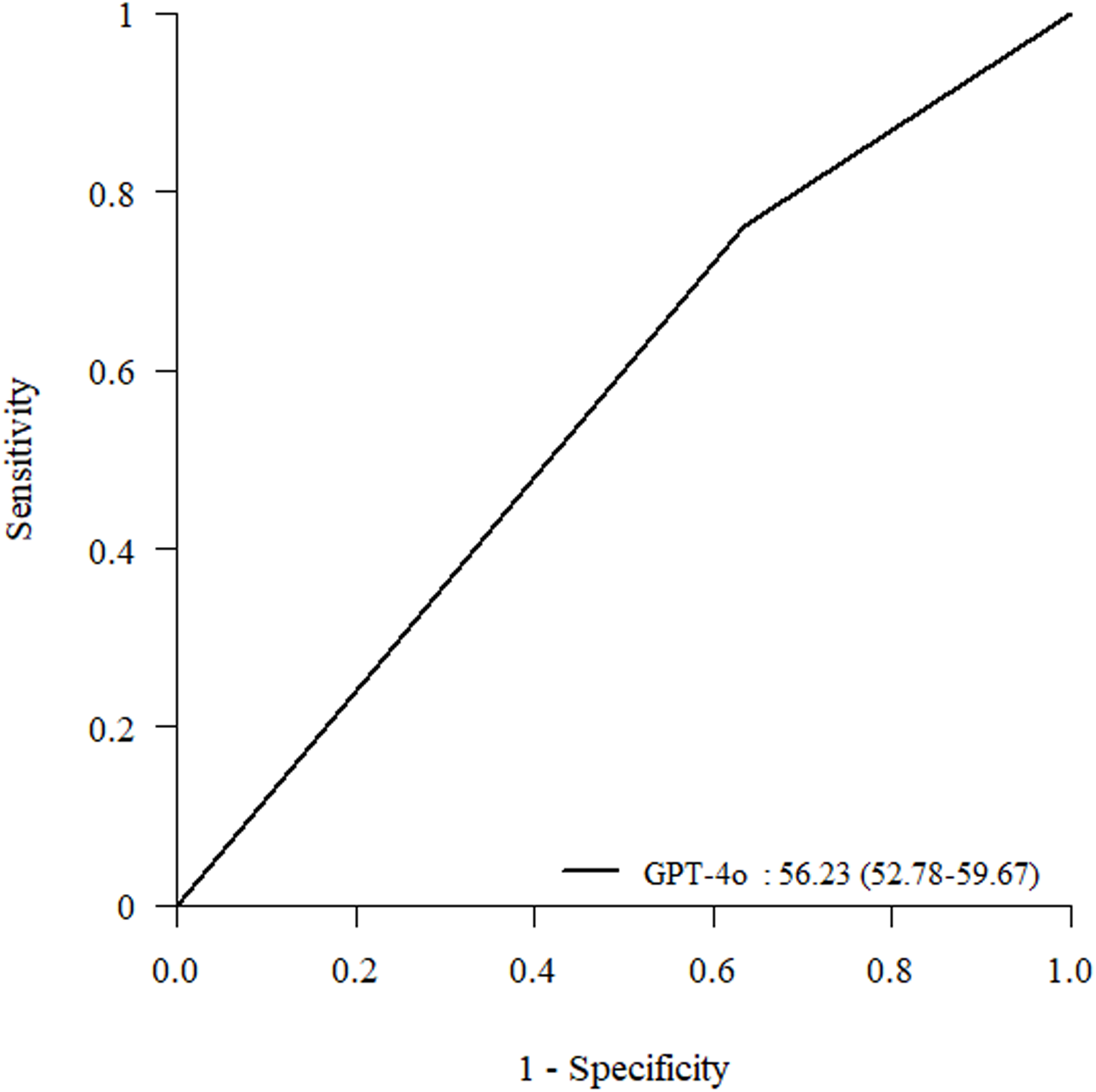
Diagnositc performance of GPT-4o and ECG Buddy. A: ChatGPT-4o, B: ECG Buddy

The dedicated ECG AI software ECG Buddy exhibited highly accurate MI classification across the STEMI, ACS, and MyoInj markers. The AUC for STEMI was 98.87% (95% CI: 98.30– 99.43), for ACS was 98.78% (95% CI: 98.05–99.50), and for MyoInj was 98.88% (95% CI: 98.24–99.51). Using the STEMI biomarker, ECG Buddy achieved the best accuracy of 96.98% (95% CI: 95.67–97.99) with a sensitivity of 96.65% (95% CI: 93.51–98.54), specificity of 97.10% (95% CI: 95.55–98.22), and F1-score of 94.27% (95% CI: 91.86–96.28). DeLong’s test confirmed that ChatGPT (AUC: 53.63%) performed significantly worse than ECG Buddy across all biomarkers (all P < .001) (Figure 3B and Table 1).

### Qualitative Assessment of Diagnostic Explanations of ChatGPT

Upon review, 17.5% (7/40) of cases lacked meaningful diagnostic explanations, with ChatGPT either failing to generate a response or providing vague and nonspecific statements. In 40% (16/40) of cases, ChatGPT provided entirely incorrect interpretations that were inconsistent with ECG morphology, either describing abnormalities not actually present or failing to identify clearly evident abnormalities. Another 35% (14/40) of cases contained partially correct interpretations. Only 7.5% (3/40) of cases demonstrated fully accurate descriptions of ECG waveform morphology, correctly identifying abnormalities in relevant leads consistent with established diagnostic criteria.

## Discussion

This study directly compared ChatGPT, a general-purpose multimodal LLM, with ECG Buddy, a specialized deep-learning tool for ECG analysis. While ECG Buddy achieved high accuracy in detecting MI from ECG images, ChatGPT’s performance was significantly inferior. The considerable performance gap between the dedicated ECG Buddy and ChatGPT underscores a fundamental difference in their architecture and training methodologies. ChatGPT is primarily optimized for textual understanding and general visual recognition tasks and lacks the specific training necessary for detailed ECG waveform interpretation. As a result, it may generate contextually plausible yet inaacurate responses, which could lead to potentially dangerous diagnostic errors if relied upon in clinical settings. Moreover, the performance of ChatGPT is highly sensitive to prompt design and the specific model version used, resulting in inconsistent outcomes. Clinical studies have reported that while ChatGPT performs moderately well in common clinical cases, it deviates significantly from evaluations in critical scenarios [10–12].Conversely, ECG Buddy is optimized through targeted domain-specific training, resulting in superior performance and reliability compared to ChatGPT.

The findings of this study indicate that current general-purpose multimodal LLM architectures may primarily rely on textual annotations or explicit labels rather than on directly analyzing waveform patterns. LLMs have potential as adjunct tools for medical consultation due to their strong language understanding capabilities; however, relying on the analysis of specialized AI models for critical diagnostic decisions is advisable. The advancement of LLMs has expanded the applications of medical AI; however, domain-specific models remain indispensable, as their fundamental limitations compromise clinical utility and reproducibility.

This study has some limitations. First, the study was conducted retrospectively using a publicly available ECG dataset, which lacked detailed clinical context or patient demographic information. The absence of comprehensive clinical data may limit the generalizability of these findings to diverse patient populations or different clinical settings. Second, qualitative assessments of the diagnostic rationales of ChatGPT were performed by a single emergency physician. While providing useful insight, interpretations by multiple clinicians across various specialties might yield different assessments of diagnostic appropriateness or accuracy. Lastly, this study evaluated a single general-purpose multimodal LLM (ChatGPT). The findings may not apply uniformly across other LLMs or future iterations with improved multimodal capabilities. Therefore, continued evaluation of newer models or subsequent versions is warranted.

Our findings are consistent with prior research highlighting that ECG-specialized AI models regularly outperform general-purpose models. Previous studies have demonstrated that deep learning models trained extensively on ECG-specific datasets accurately detect subtle waveform changes indicative of asymptomatic ventricular dysfunction and cirrhosis and can infer patient characteristics such as sex and age [22–24]. Similar to these specialized ECG models, ECG Buddy undergoes targeted optimization tailored specifically to ECGimages, ensuring consistent predictive performance and stable error margins essential for clinical reliability. Notably, Users only need to provide an ECG image or screenshot; both the smartphone and desktop versions feature an intuitive interface that requires no extra training or expertise. In addition to MI, ECG Buddy demonstrates robust diagnostic capabilities across diverse cardiac conditions, including STEMI, hyperkalemia, and RV dysfunction, validating its efficacy in various clinical settings [15–18]. Moreover, it has even outperformed human experts in diagnosing STEMI and hyperkalemia.

## Conclusions

To the best of our knowledge, this study provides the first direct comparative assessment between ChatGPT and ECG Buddy for detecting MI from ECG images. Our findings reveal that, despite the accelerating use of LLM-based AI, current LLMs do not meet the clinical performance and accuracy requirements for ECG interpretation. Herein, the ability of a general-purpose multimodal LLM (ChatGPT) to detect ECG abnormalities fell significantly short of that achieved by board-certified emergency physicians, rendering it insufficient for use in interpreting critical ECG readings in clinical practice. In contrast, the specialized ECG AI tool (ECG Buddy), which has been thoroughly trained and validated, achieved high diagnostic accuracy and reproducibility, confirming its utility in clinical settings. These results, consistent with those of several other studies, underscore the superiority of medical domain-specific AI programs over general-purpose LLMs for ECG analysis and interpretation and emphasize the importance of specialized models in the field of medical AI development.

## Data Availability

All data produced in the present study are available upon reasonable request to the authors

https://data.mendeley.com/datasets/gwbz3fsgp8/2

---

**Textbox 1.**
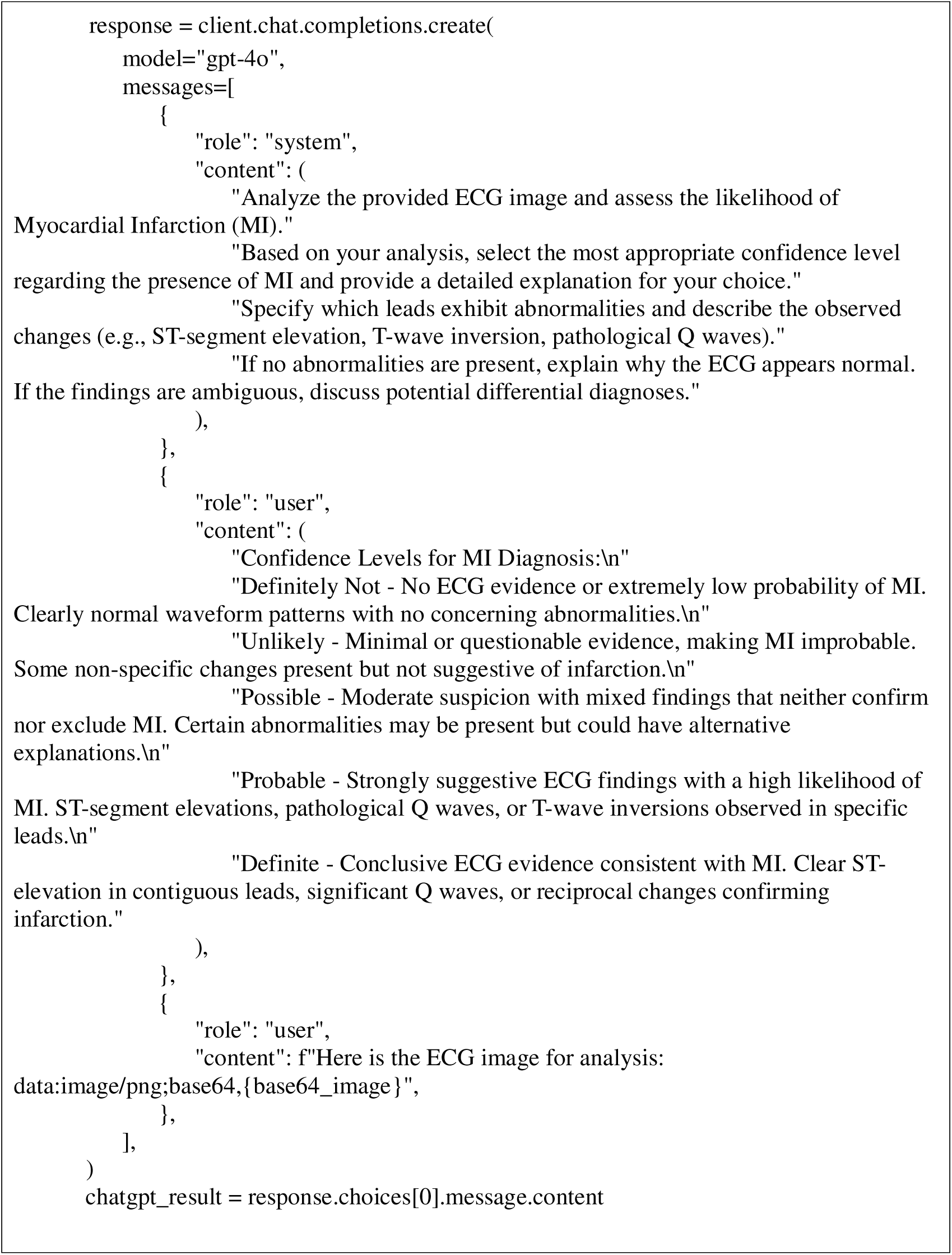
Generative Pre-trained Transformer (GPT)-4o prompt design.

**Table 2.** Performance Summary.

|  | Chat Generative Pre-trained Transformer | ECG Buddy |  |  |
| --- | --- | --- | --- | --- |
|  |  | ST-elevation myocardial infarction | Acute coronary syndrome | Myocardial injury |
| Sensitivity | 36.40 (30.30–42.85) | 96.65 (93.50–98.54) | 96.65 (93.51–98.54) | 96.65 (93.51–98.54) |
| Specificity | 76.20 (72.84–79.33) | 97.10 (95.55–98.22) | 96.66 (95.03–97.87) | 97.24 (95.73–98.33) |
| PPV | 34.66 (28.79–40.90) | 92.03 (87.96–95.07) | 90.94 (86.72–94.17) | 92.40 (88.39–95.36) |
| NPV | 77.55 (74.21–80.64) | 98.82 (97.68–99.49) | 98.81 (97.67–99.49) | 98.82 (97.69–99.49) |
| AUROC | 57.34 (53.44–61.24) | 98.87 (98.30–99.43) | 98.78 (98.05–99.50) | 98.88 (98.24–99.51) |
| Accuracy | 65.95 (62.80–69.00) | 96.98 (95.67–97.99) | 96.66 (95.29–97.72) | 97.09 (95.79–98.07) |

Data are expressed as values (95% confidence interval). ECG, electrocardiogram; AUROC, area under the receiver operating curve; PPV, positive predictive value; NPV, negative predictive value

## Author contributions

1. Conceptualization: Keehyuck Lee, Dongbum Suh, Joonghee Kim, Youngjin Cho
2. Data curation: Haemin Lee, Sooyoung Yoo
3. Formal analysis: Haemin Lee, Sooyoung Yoo
4. Funding acquisition: Joonghee Kim, Youngjin Cho
5. Investigation: Haemin Lee, Sooyoung Yoo
6. Methodology: Haemin Lee, Joonghee Kim
7. Project administration: Keehyuck Lee, Dongbum Suh
8. Resources: Joonghee Kim, Youngjin Cho
9. Supervision: Keehyuck Lee, Dongbum Suh
10. Validation: Haemin Lee
11. Visualization: Haemin Lee
12. Writing – Haemin Lee, Joonghee Kim
13. Writing – review & editing: Keehyuck Lee, Dongbum Suh, Sooyoung Yoo
14. Approval of final manuscript: all authors

## Acknowledgments

This research was partly supported by a grant from the Technological Innovation Research & Development Program (SCALEUP TIPS), funded by the Ministry of SMEs and Startups (grant number: RS-2024-00415492), and the Medical AI Clinic Program through the National IT Industry Promotion Agency, funded by the Ministry of Science Information Communication and Technology (grant number: H0904-24-1002).

